# Thrombectomy in addition to thrombolysis for medium distal vessel occlusions - results from the SITS registry

**DOI:** 10.1101/2025.09.03.25335063

**Authors:** Boris Keselman, Michael Mazya, Daniel Strbian, Ana Paiva Nunes, Andrea Naldi, Danilo Toni, Marco Petruzzellis, Giovanni Frisullo, Dalius Jatuzis, Silvia Strumia, Niaz Ahmed, Tiago Moreira

**Author notes:** **Corresponding author** Boris Keselman, MD, PhD, Karolinska Stroke Research Unit, Carolina Tower Hotel, floor 4, Karolinska University Hospital – Solna, Stockholm Sweden.

## Abstract

**Background:** Endovascular thrombectomy (EVT) is standard of care for acute ischemic stroke (AIS) caused by large vessel occlusion. Medium distal vessel occlusions (MDVO) account for 25-40% AIS cases, but recanalization rates with intravenous thrombolysis (IVT) are often below 50%. Studies have shown mixed results comparing EVT with best medical management in different MDVO populations. This study aims to show if EVT in addition to IVT is associated with clinical benefits or harm in patients with MDVO.

**Methods:** We performed an observational study of patients in the Safe Implementation of Treatments of Stroke International Stroke Registry (SITS-ISTR) 2016–23, treated with IVT or IVT+EVT for occlusion of the ACA, PCA or distal MCA (M3 and more distal). Outcomes were modified Rankin Scale score (mRS), death at 3 months, and post-treatment hemorrhage. Inverse probability treatment weighting and binomial logistic regression was performed due to baseline imbalances (age, NIHSS and occlusion site).

**Results:** Of 2198 included patients, 1903 (87%) received IVT, and 295 (13%) IVT+EVT. IVT+EVT patients were younger (73 vs 75) and had higher median NIHSS: 10 (IQR 6-15) vs 8 (5-12), p<0.001. The most common occlusion site was PCA in the IVT+EVT group, (n=179, 60.7%), and distal MCA (n=1140, 59.9%) in the IVT group. Rates of symptomatic intracerebral hemorrhage (SICH) were higher in the IVT+EVT group (SICH NINDS: 7.6% vs 3.5%, p=0.003; SICH ECASS-II: 5.4% vs 2.4%, p=0.011). After adjustment, IVT+EVT was associated with worse outcomes compared to IVT alone (aOR, 95% CI); mRS 0-1: 0.63 (0.44-0.90), mRS 0-2: 0.64 (0.44-0.91) and death: 2.03 (1.26-3.27).

**Conclusions:** IVT+EVT for MDVO was associated with worse outcomes compared to IVT alone. Results should be interpreted with caution due to retrospective observational design, warranting further randomized studies, but are in line with recently published RCTs

## Introduction

Endovascular thrombectomy (EVT) has been well established in randomized controlled trials as treatment in acute ischemic stroke caused by proximal large vessel occlusion (PLVO), both in the anterior and posterior circulation.^1,2^ Medium distal vessel occlusions (MDVO), however, account for around 40% of ischemic stroke cases and treatment with intravenous thrombolysis is often insufficient for recanalization.^3^ Recently, two randomized controlled trials have shown no benefit of EVT in addition to best medical therapy (BMT). Despite methodological differences in inclusion criteria (dominant/non-dominant M2, A1/P1 segments) as well as time window (12h vs 24h), neither trial demonstrated statistically significant benefit nor harm in patients treated with EVT.^4,5^ The majority of patients had an MCA occlusion and the most common site was M2 for both trials. Previously, several observational studies have shown conflicting results for EVT in MDVO. Three recent studies on isolated posterior cerebral artery (PCA) occlusion have compared EVT to best medical treatment (BMT), that may or may not include IVT in either group. Two studies have shown an association with better functional outcomes for those treated with EVT without an increased risk for intracerebral hemorrhage.^6,7^ However, the third study, with a sub-analysis comparing EVT +/- IVT vs IVT alone, showed a higher mortality and risk of SICH in the EVT group, without a difference in mRS.^8,9^ In comparisons of EVT with BMT for MDVO including the anterior circulation, results have similarly varied between lower risk of SICH without higher odds of better functional outcome, or better functional outcome without any differences in risk of SICH or death.^10,11^ Further randomized controlled trials comparing EVT to BMT in MDVO are ongoing.^12,13^ Additionally, one was recently terminated following a lack of efficacy of EVT for MDVO.^14^ The aim of this study is to compare the safety and outcomes of treatment with IVT or IVT and EVT in distal MDVO stroke, excluding M2 occlusions.

## Methods

The corresponding author had full access to the data in the study and takes full responsibility for its integrity and the data analysis. Access to the anonymized data for this study will be available from the corresponding author upon reasonable request from qualified researchers, contingent on approval by the SITS Scientific Committee. All patients treated with IVT recorded in the SITS International Stroke Treatment Register (SITS-ISTR) between 2016 and 2023, with available occlusion data from CTA or MRA imaging were considered for this study. MDVO was defined as any isolated occlusion in the ACA, PCA or distal segments of the MCA (M3 or more distal). Patients treated only with IVT formed the IVT group, whereas those treated with both IVT and EVT formed the IVT+EVT group. Treatment decisions were made at the discretion of physicians at the participating centers.

The SITS-ISTR is an ongoing, prospective, internet-based, academic-driven, multinational, observational monitoring register for clinical centers using thrombolysis for the treatment of acute ischemic stroke. The methodology of the SITS-ISTR, including procedures for data collection and management, patient identification and verification of source data, has been described previously.^15–17^

We used collected data on baseline and demographic characteristics, stroke severity per the standard 15-item, 42-point NIHSS, time logistics, medication history, and imaging data on admission and follow-up, as well as modified Rankin Scale (mRS) at three months follow-up.

### Outcomes

Efficacy outcomes were the full range of the mRS, as well as the standard dichotomizations functional independence (mRS 0-2) and excellent functional outcome (mRS 0-1). Safety outcomes were any posttreatment intracerebral hemorrhage, subdivided into hemorrhagic infarction (HI), parenchymal hematoma (PH), as well as SICH per the SITS-MOST, ECASS II and NINDS definitions.^18^ Death at three months was also assessed as a safety outcome. All SICH events were adjudicated centrally by the SITS International Coordination Office based on submitted clinical and imaging reports; images were not available for review.

### Statistical analysis

We performed descriptive statistical analyses for baseline demographic, clinical and imaging data, as well as outcomes, comparing the IVT and IVT+EVT groups. For continuous variables, median and interquartile range values were calculated. For categorical variables, percentage proportions were calculated by dividing the number of events by the total number of patients, excluding missing or unknown cases, according to standard SITS methodology.^19,20^ Calculations of significance of difference between proportions were performed using the Pearson χ^2^ method. P values of <0.05 (two-tailed) were considered significant.

Furthermore, outcome analysis was performed with propensity score matching (PSM) analysis to account for clinically and statistically significant baseline differences between the two treatment groups. Age, sex, minor stroke (NIHSS < 6) and occlusion site (MCA, ACA or PCA) were used as variables. The propensity score for treatment with IVT+EVT was calculated for each patient and a nearest neighbor algorithm was used with a ratio of 1:2 and a caliper of 0.1 to compare the two groups. Absolute standardized mean differences were calculated before and after matching to ensure balance in the PSM model (see supplemental figure I in the online supplement at https://www.ahajournals.org/journal/str. Binomial logistic regression was performed for the outcomes after matching. A sensitivity analysis for the outcomes was performed using binomial logistic regression after PSM with pre-stroke mRS and the full range of the NIHSS added as additional covariates. Statistical analyses were performed in R 4.3.1 (https://www.R-project.org/). Results are reported according to STROBE.^21^

### Ethics approval and data monitoring

Ethics approval was obtained from the Stockholm Regional Ethics Committee for this project as part of the SITS-MOST (Safe Implementation of Thrombolysis in Stroke Monitoring Study) II framework. The SITS International Coordination Office monitored the SITS-ISTR data online and checked individual patient data monthly to identify errors or inconsistencies.

## Results

We identified 2198 patients with a MDVO stroke, of which 1903 were treated with IVT and 295 were treated with IVT + EVT between 2016 and 2023, see figure I. Table I shows a comparison of characteristics between the two treatment groups. Patients in the IVT + EVT group were more commonly male (60.3% vs 51.2%), younger (median 73 vs 75 years), had higher baseline stroke severity (median NIHSS 10 vs 8), as well as different distributions of occluded arteries (PCA: 60.7% vs 30.4%; distal MCA: 24.1% vs 59.9%; ACA: 15.3% vs 9.7%), all p<0.001. There was also a higher proportion of patients with baseline mRS score of 0-1 before stroke in the IVT + EVT group (91.8% vs 83.0%, p < 0.001). Other baseline characteristics were similar between the two groups.

**Figure I.**
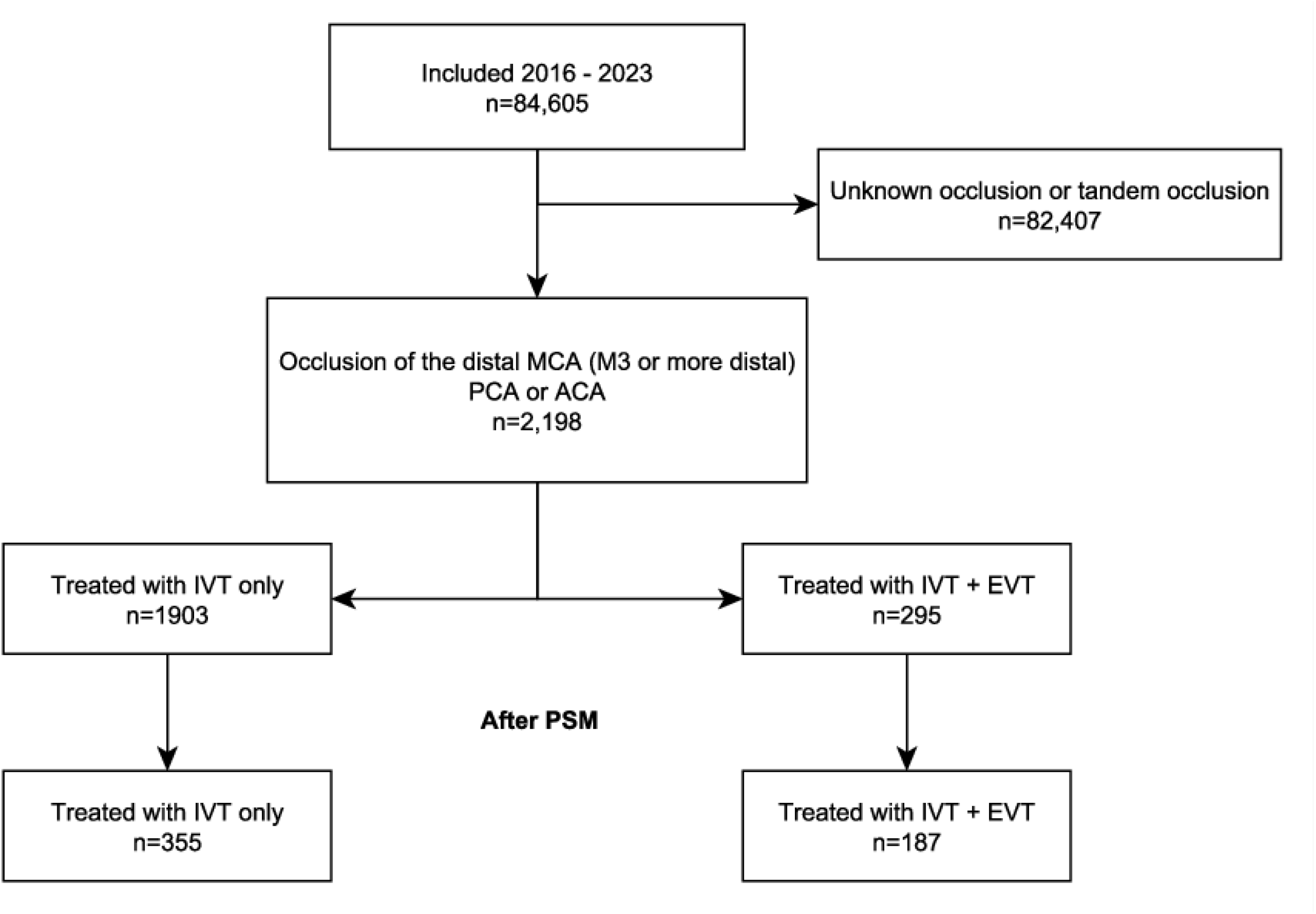
Patient flow diagram. IVT: intravenous thrombolysis; LVO: large vessel occlusion; MCA: middle cerebral artery; PCA: posterior cerebral artery; ACA: anterior cerebral artery; EVT: endovascular thrombectomy; PSM: propensity score matching

**Table I.** Univariate comparison of characteristics between treatment groups.

|  | IVT (n=1903) | IVT+EVT (n=295) | p value |
| --- | --- | --- | --- |
| Age, median, (IQR) | 75 (67-82) | 73 (61-79) | <0.001 |
| Sex, female, (%) | 928 (48.8) | 117 (39.7) | 0.004 |
| Pre-stroke mRS 0-1 (%) | 1518 (83.0) | 256 (91.8) | <0.001 |
| NIHSS (median, IQR) | 8 (5-12) | 10 (6-15) | <0.001 |
| NIHSS > 6 (%) | 1097 (58.4) | 214 (74.6) | <0.001 |
| Comorbidities (%) |  |  |  |
| Hypertension | 1411 (74.6) | 199 (68.2) | 0.023 |
| Smoking (current/previous) | 446 (24.8) | 69 (25.1) | 0.990 |
| Diabetes | 435 (23.0) | 66 (22.5) | 0.912 |
| Hyperlipidemia | 627 (33.8) | 92 (31.8) | 0.554 |
| Atrial fibrillation | 147 (11.5) | 16 (8.2) | 0.215 |
| Congestive heart failure | 149 (7.9) | 16 (5.5) | 0.182 |
| Medications (%) |  |  |  |
| Antiplatelet therapy | 506 (37.8) | 60 (30.9) | 0.075 |
| Oral anticoagulants | 97 (5.4) | 12 (4.1) | 0.444 |
| Systolic blood pressure (mean, SD) | 155 (25) | 153 (25) | 0.149 |
| Diastolic blood pressure (mean, SD) | 84 (15) | 84 (15) | 0.475 |
| Occlusion site |  |  | <0.001 |
| MCA (M3 or more distal) | 1140 (59.9) | 71 (24.1) |  |
| PCA | 578 (30.4) | 179 (60.7) |  |
| ACA | 185 (9.7) | 45 (15.3) |  |
| Onset to hospital, min (median, IQR) | 89 (62-132) | 87 (57-129) | 0.285 |
| Onset to puncture, min (median, IQR) | NA | 221 (175-313) | NA |

Table legend. IVT: intravenous thrombolysis; EVT: endovascular thrombectomy; mRS: modified Rankin Scale; NIHSS: National Institutes of Health Stroke Scale; MCA: Middle Cerebral Artery; PCA: Posterior Cerebral Artery; ACA: Anterior Cerebral Artery

Table II shows unadjusted comparisons of outcomes between the two groups as well as outcomes after PSM. At three months follow-up, fewer patients treated with IVT+EVT achieved functional independence (54.9% vs 69.0%) or excellent functional outcome (65.6% vs 77.0%), both p<0.001. More patients in the IVT+EVT group died within 3 months, 20.9% vs 13.6%, p=0.008. Figure II shows the full distribution of mRS scores at three months in the two treatment groups. For acute bleeding complications, more patients with IVT+EVT had a hemorrhagic infarction (7.3% vs 11.6%, p=0.020), as well as SICH per NINDS and ECASS II definitions. The rates of parenchymal hematoma and SICH per SITS-MOST were similar in the two groups.

**Figure II.**
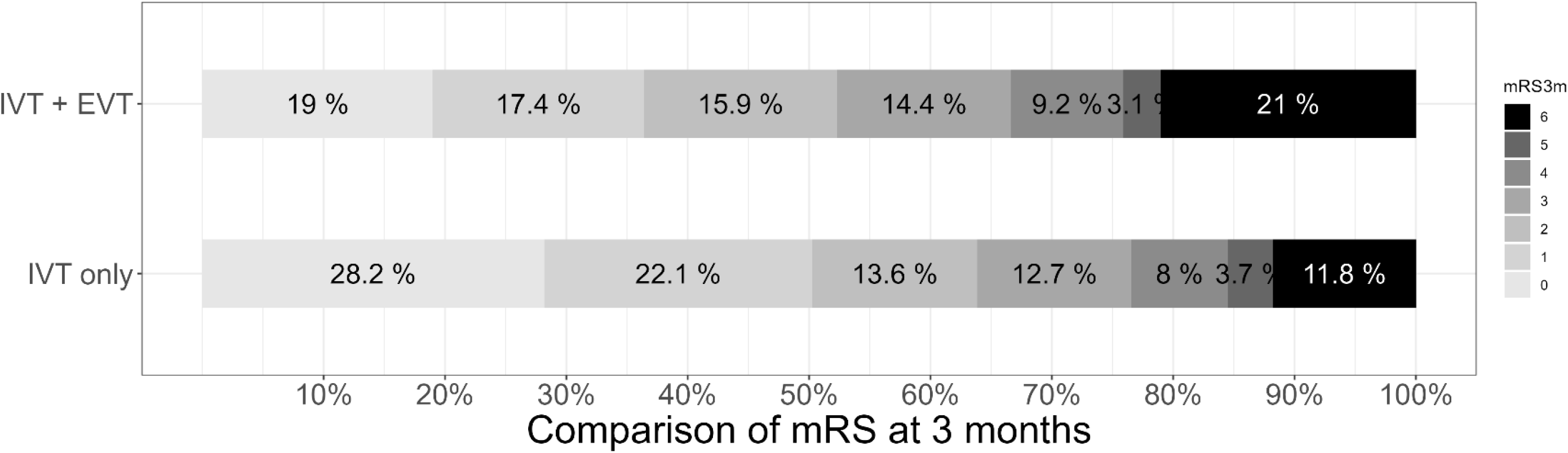
distribution of mRS at three months follow up IVT: intravenous thrombolysis; EVT: endovascular thrombectomy; mRS: modified Rankin Scale score.

**Table II.**
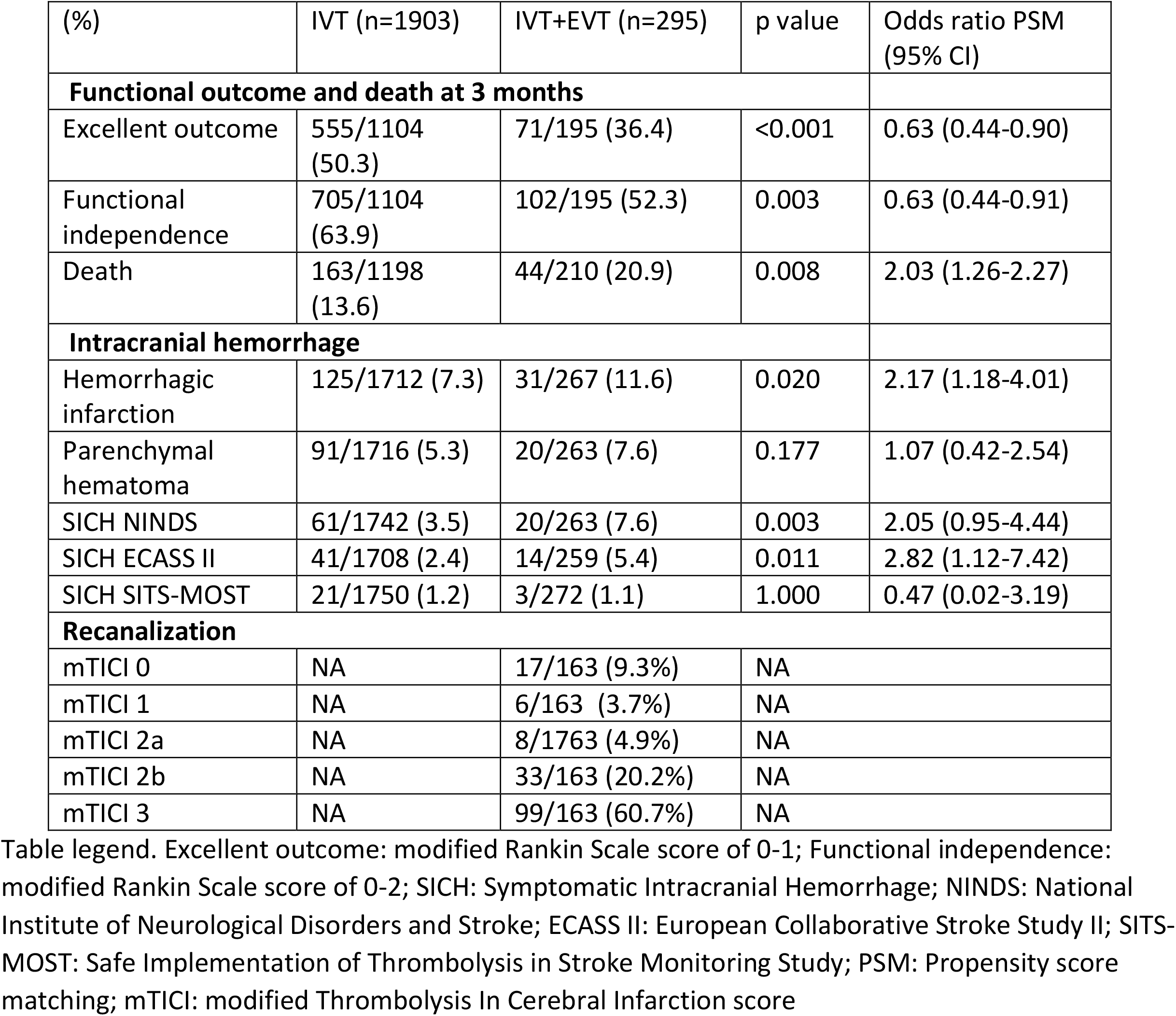
outcomes.

After PSM analysis, the effective sample size was 187 in the IVT+EVT group and 355 in the IVT only group (see figure I). Baseline characteristics after PSM are presented table I in the online supplement (please see https://www.ahajournals.org/journal/str) with no statistically significant differences in age, sex, proportion of patients with minor stroke and occlusion site. Statistically significant differences remained in the proportion of patients with pre-stroke mRS 0-1 (IVT: 82.9% vs IVT + EVT: 93.5%) as well as the full range of NIHSS score at baseline (IVT: 8, IQR 5-11 vs IVT + EVT: 10, IQR 6-15). Outcome results after PSM are presented in table II, showing lower odds ratios of excellent outcome (0.63, 95% CI: 0.44-0.90), functional independence (0.63, 95% CI: 0.44-0.91) and a higher odds ratio of death (2.03, 95% CI: 1.26-2.27) at three months for the IVT+EVT group. For bleeding outcomes, there were higher odds ratios of hemorrhagic infarction (2.17, 95% CI: 1.18-4.01) and SICH per ECASS II (2.82, 95% CI: 1.12-7.42), but not for parenchymal hematoma, SICH per SITS-MOST or SICH per NINDS. The sensitivity analysis using multivariate binomial logistic regression after PSM showed similar results, reported in table III of the online supplement (please see https://www.ahajournals.org/journal/str).

## Discussion

This observational study, performed on a large, international stroke-registry, found that treatment of patients with MDVO stroke using IVT + EVT was associated with worse outcomes compared to IVT. The higher mRS scores in the IVT + EVT group may be due to a higher rate of bleeding complications occurring in the acute in-hospital period. These results further strengthen the findings of two recently published randomized controlled trials showing no benefit of added EVT compared to best medical therapy alone and highlight the need to evaluate which patients are appropriate for endovascular treatment.^4,5^

Since the first positive trials in 2015, several studies have widened the indications for EVT, opening possibilities for treatment in previously excluded groups such as those with symptom onset > 6 hours, basilar artery occlusion and large infarctions on baseline imaging.^2,22–25^ Meanwhile, other studies have shown that there may be a limit to the effectiveness of EVT in certain populations, such as those with minor stroke.^26,27^ In light of these findings, routine use of EVT has expanded dramatically, impacting the inclusion of patients in randomized trials. When compared to the patients included in the HERMES collaboration, patients included in the DISTAL, ESCAPE-MEVO and DISCOUNT trials were older (median age 74-77 vs 68), had lower stroke severity (median NIHSS 6-8 vs 17) and were less often treated with IVT (ranging from 57 to 71% vs 85%), as well as being randomized at later time-points (234-261 vs 196 min).^1,4,5^ Our study, with slightly higher stroke severity, may be more representative of the patients treated for MDVO in routine clinical practice. One explanation for the worse outcomes reported in our study compared to the recent randomized trials may be the exclusion of M2 occlusions, which may have a more favorable response to treatment with EVT as shown previously.^28^

While previous observational studies have compared EVT to best medical therapy for MDVO, this is, to our knowledge, the only study where the entire cohort has been treated with IVT. The added benefit of this methodology is the possibility to compare the addition of EVT for patients with active reperfusion treatment in both groups. We used PSM to increase the comparability of the treatment groups, accounting for clinically relevant baseline differences as well as a sensitivity analysis using multivariate logistic regression after PSM including pre-stroke mRS and the full range of the NIHSS, which both showed similar results.

This study has several limitations, including those inherent in a retrospective, observational registry-based design such as risk of bias and missing data. Information on mRS at three months was missing in 40% of patients (34% for IVT+EVT and 41% in IVT). Data availability was over 90% for acute bleeding complications in both groups. Comparability of the two treatment groups is complicated due to confounding by indication, as well as the possibility of different levels of care available in a thrombectomy capable stroke center vs a center with only thrombolysis available for reperfusion. Furthermore, patients treated with IVT+EVT had more severe strokes (median NIHSS 10 vs 8), which may have been offset by other favorable baseline characteristics: a higher proportion of pre-stroke mRS 0-1 and younger age. However, after PSM, differences in both acute bleeding outcomes as well as functional outcomes remained statistically significant in favor of treatment with IVT only.

## Conclusion

The results from this observational, retrospective study show that treatment with IVT+EVT for MDVO stroke is associated with worse functional outcomes compared to treatment with IVT alone. This could be explained with a higher proportion of patients with acute intracranial bleeding complications in this group. Future randomized trials must be carefully designed to improve patient selection but also to improve selection of adequate endovascular devices and techniques since the risk benefit ratio of IVT+EVT in MDVO is different compared to LVO.

## Data Availability

The corresponding author had full access to the data in the study and takes full responsibility for its integrity and the data analysis. Access to the anonymized data for this study will be available from the corresponding author upon reasonable request from qualified researchers, contingent on approval by the SITS Scientific Committee.

## Declarations

Conflicting interests: The Authors declare that there is no conflict of interest

## Non-standard Abbreviations and Acronyms

SITS-ISTR: Safe Implementation of Treatments in Stroke - International Stroke Registry
AIS: Acute ischemic stroke
EVT: Endovascular thrombectomy
IVT: Intravenous thrombolysis
BMT: Best medical therapy
PLVO: Proximal large vessel occlusion
MDVO: Distal medium vessel occlusion
mRS: Modified Rankin Scale
NIHSS: National Institute of Health Stroke Scale
SICH: Symptomatic Intracranial Hemorrhage

## References

1. Goyal M, Menon BK, van Zwam WH, et al. Endovascular thrombectomy after large-vessel ischaemic stroke: a meta-analysis of individual patient data from five randomised trials. Lancet Lond Engl 2016; 387: 1723–1731.

2. Jovin TG, Li C, Wu L, et al. Trial of Thrombectomy 6 to 24 Hours after Stroke Due to Basilar-Artery Occlusion. N Engl J Med 2022; 387: 1373–1384.

3. Saver JL, Chapot R, Agid R, et al. Thrombectomy for Distal, Medium Vessel Occlusions: A Consensus Statement on Present Knowledge and Promising Directions. Stroke 2020; 51: 2872–2884.

4. Psychogios M, Brehm A, Ribo M, et al. Endovascular Treatment for Stroke Due to Occlusion of Medium or Distal Vessels. N Engl J Med; 0. DOI: 10.1056/NEJMoa2408954.

5. Goyal M, Ospel JM, Ganesh A, et al. Endovascular Treatment of Stroke Due to Medium-Vessel Occlusion. N Engl J Med; 0. DOI: 10.1056/NEJMoa2411668.

6. Dicpinigaitis AJ, Syed SA, Al-Mufti J, et al. Endovascular thrombectomy for treatment of isolated posterior cerebral artery occlusion: a real-world analysis of hospitalizations in the United States. Acta Neurochir (Wien) 2024; 166: 191.

7. Meyer L, Stracke CP, Jungi N, et al. Thrombectomy for Primary Distal Posterior Cerebral Artery Occlusion Stroke: The TOPMOST Study. JAMA Neurol 2021; 78: 434– 444.

8. Nguyen TN, Qureshi MM, Strambo D, et al. Endovascular Versus Medical Management of Posterior Cerebral Artery Occlusion Stroke: The PLATO Study. Stroke 2023; 54: 1708–1717.

9. Räty S, Nguyen TN, Nagel S, et al. Endovascular Thrombectomy Versus Intravenous Thrombolysis of Posterior Cerebral Artery Occlusion Stroke. J Stroke 2024; 26: 290– 299.

10. Saber H, Desai SM, Haussen D, et al. Endovascular Therapy vs Medical Management for Patients With Acute Stroke With Medium Vessel Occlusion in the Anterior Circulation. JAMA Netw Open 2022; 5: e2238154.

11. Mohammaden MH, Souza Viana L, Abdelhamid H, et al. Endovascular Versus Medical Management in Distal Medium Vessel Occlusion Stroke: The DUSK Study. Stroke 2024; 55: 1489–1497.

12. Hu W. Evaluation of Endovascular Treatment in Acute Intracranial Distal Medium Vessel Occlusion Stroke - a Multicenter, Randomized Controlled, Clinical Trial. Clinical Trial Registration NCT06146790, clinicaltrials.gov, https://clinicaltrials.gov/study/NCT06146790 (31 August 2024, accessed 22 April 2025).

13. ANZCTR - Registration, https://www.anzctr.org.au/Trial/Registration/TrialReview.aspx?id=384551 (accessed 22 April 2025).

14. Assistance Publique - Hôpitaux de Paris. Evaluation of Mechanical Thrombectomy in Acute Ischemic Stroke Related to a Distal Arterial Occlusion: a Randomized Controlled Trial. Clinical Trial Registration NCT05030142, clinicaltrials.gov, https://clinicaltrials.gov/study/NCT05030142 (14 April 2023, accessed 1 January 2024).

15. Mazya MV, Cooray C, Lees KR, et al. Minor stroke due to large artery occlusion. When is intravenous thrombolysis not enough? Results from the SITS International Stroke Thrombolysis Register. Eur Stroke J 2018; 3: 29–38.

16. Wahlgren N, Ahmed N, Dávalos A, et al. Thrombolysis with alteplase for acute ischaemic stroke in the Safe Implementation of Thrombolysis in Stroke-Monitoring Study (SITS-MOST): an observational study. Lancet Lond Engl 2007; 369: 275–282.

17. Keselman B, Cooray C, Vanhooren G, et al. Intravenous thrombolysis in stroke mimics: results from the SITS International Stroke Thrombolysis Register. Eur J Neurol; 0. DOI: 10.1111/ene.13944.

18. von Kummer Rüdiger, Broderick Joseph P., Campbell Bruce C.V., et al. The Heidelberg Bleeding Classification. Stroke 2015; 46: 2981–2986.

19. Cooray Charith, Mazya Michael, Mikulik Robert, et al. Safety and Outcome of Intravenous Thrombolysis in Stroke Patients on Prophylactic Doses of Low Molecular Weight Heparins at Stroke Onset. Stroke 2019; 50: 1149–1155.

20. Mazya Michael V., Ahmed Niaz, Azevedo Elsa, et al. Impact of Transcranial Doppler Ultrasound on Logistics and Outcomes in Stroke Thrombolysis. Stroke 2018; 49: 1695– 1700.

21. Elm E von, Altman DG, Egger M, et al. Strengthening the reporting of observational studies in epidemiology (STROBE) statement: guidelines for reporting observational studies. BMJ 2007; 335: 806–808.

22. Nogueira RG, Jadhav AP, Haussen DC, et al. Thrombectomy 6 to 24 Hours after Stroke with a Mismatch between Deficit and Infarct. N Engl J Med 2018; 378: 11–21.

23. Albers GW, Marks MP, Kemp S, et al. Thrombectomy for Stroke at 6 to 16 Hours with Selection by Perfusion Imaging. N Engl J Med 2018; 378: 708–718.

24. Sarraj A, Hassan AE, Abraham MG, et al. Trial of Endovascular Thrombectomy for Large Ischemic Strokes. N Engl J Med 2023; 388: 1259–1271.

25. Bendszus M, Fiehler J, Subtil F, et al. Endovascular thrombectomy for acute ischaemic stroke with established large infarct: multicentre, open-label, randomised trial. Lancet Lond Engl 2023; 402: 1753–1763.

26. Safouris A, Palaiodimou L, Nardai S, et al. Medical Management Versus Endovascular Treatment for Large-Vessel Occlusion Anterior Circulation Stroke With Low NIHSS. Stroke 2023; 54: 2265–2275.

27. Schwarz G, Bonato S, Lanfranconi S, et al. Intravenous thrombolysis + endovascular thrombectomy versus thrombolysis alone in large vessel occlusion mild stroke: a propensity score matched analysis. Eur J Neurol 2023; 30: 1312–1319.

28. Menon BK, Hill MD, Davalos A, et al. Efficacy of endovascular thrombectomy in patients with M2 segment middle cerebral artery occlusions: meta-analysis of data from the HERMES Collaboration. J Neurointerventional Surg 2019; 11: 1065–1069.

